# Stay or go? Outcomes of lower limb arthroplasty in patients travelling away from home for surgery: A cross-sectional analysis of the AOANJRR comparing patient residence and hospital remoteness

**DOI:** 10.1101/2024.08.25.24312205

**Authors:** Corey Scholes, Carl Holder, Christopher Vertullo, Matthew Broadhead

## Abstract

**Purpose:** The relationship between remoteness of patient residence and post-surgical outcomes, such as early implant revision, has yet to be examined. The aim of this study was to assess whether the incidence of all-cause revision at up to 2 years following primary hip or knee total joint arthroplasty varies with the remoteness of a person’s place of residence at the time of the primary procedure.

**Methods:** An analysis was performed of data from the Australian Orthopaedic Association National Joint Replacement Registry (AOANJRR) from 1 Sept 1999 to 31 Dec 2022. The Modified Monash Model (2015) of remoteness classification was used to categorise patient residence and hospital location into metro-regional (MM 1-2) and rural-remote (MM 3-7). All-cause revision within the two-year period after surgery for primary total knee arthroplasty (TKA) and primary total hip arthroplasty (THA) for osteoarthritis as the primary diagnosis was selected as the primary outcome. A directed-acyclic graph approach was used to prioritise covariates for inclusion in a Cox proportional hazards regression model. Cumulative percent revision (CPR) rates with 95% confidence intervals was reported with hazard ratios between subgroups of residential and hospital remoteness.

**Results:** The two-year CPR for primary TKA ranged from 1.8% (95% CI 1.7 - 1.9) to 2.2% (95% CI 1.8 - 2.7). Patients residing in rural-remote areas who travelled to a metro-regional hospital displayed a significantly higher rate of revision following TKA compared to patients that were treated at a rural-remote hospital (HR: 1.11, 95% CI 1.05 - 1.18, P = 0.001) within two-year follow-up of the primary procedure. Patients residing in rural-remote areas that stayed in these areas for their operation displayed a significantly reduced revision rate compared to metro-regional patients that stayed in-area for their joint replacement (HR=0.90, 95%CI 0.85 - 0.95, P <0.001). Infection was the dominant reason for TKA revision for patients in the follow-up period. No discernible differences in revision risk were observed between patient and hospital combinations for primary total hip replacement.

**Conclusions:** Travel distance, but not remoteness of a patient’s place of residence may be associated with cumulative risk of early revision (within 2 years) of primary TKA, particularly in regional/remote patients travelling out of area, but not for patients undergoing THA. Further work linking service utilisation prior to a revision procedure is required to clarify whether differences in revision between remoteness and travel distances are due to variability in the clinical threshold for offering revision arthroplasty between regional and metropolitan surgeons or improved outcomes of the primary procedure.

## Introduction

The outcomes of regional patients who travel to capital cities for lower limb joint arthroplasty compared to those who undergo the surgery in the same geographic area remains uncertain. A recent parliamentary inquiry in New South Wales found rural, regional and remote patients have significantly poorer health outcomes, greater incidence of chronic disease and higher risk of premature death when compared to their counterparts in metropolitan areas, with inferior access to health and hospital services (Legislative Council. Portfolio Committee No. 2 – Health, 2022). In other settings, the rural-metropolitan disparity in health and mortality outcomes is of increasing concern (Cosby et al., 2019; Harrington et al., 2020). Osteoarthritis, particularly of the hip and knee, remains a global health challenge with increasing prevalence (Long et al., 2022), and a concomitant increase in burden of joint replacement surgery (Ackerman et al., 2019). A common perception is that care offered in a higher volume tertiary metropolitan centre is associated with superior surgical outcomes for lower limb arthroplasty. Although this perception is broadly accepted, differences in revision outcomes after primary lower limb arthroplasty have not been readily apparent (Alvarez et al., 2022; S. L. Brennan-Olsen et al., 2019). Nevertheless, assessment of revision risk with respect to the place of residence of patients remains lacking in the literature.

Worsening outcomes with increased distance from metropolitan centres for other conditions such as colorectal cancer in Australia have been previously reported (Ireland et al., 2017). The authors argued that access to services and treatments may not be the major drivers of disparities between metropolitan and regional areas, and may be better explained by individual patient characteristics and regional characteristics such as socioeconomic status, as well as patient age and patient sex at the time of diagnosis. Other studies have shown relationships between place of residence (rural-urban) and cancer survival in other high-income countries (Afshar et al., 2019). In contrast, one study reported that place of residence as well as other system and regional characteristics were not associated with any outcomes of interest following surgery for colorectal cancer and in fact patient characteristics such as older age, presence of comorbidities and previous emergency admissions were stronger explanatory variables (Theile et al., 2019). More relevant for arthroplasty surgery, disparities in utilisation have been observed with respect to socioeconomic position, rather than geography, within a regional setting of Australia (S. Brennan-Olsen et al., 2017) and internationally (Alvarez et al., 2022). However, the relationship between remoteness of patient residence and post-surgical outcomes has yet to be examined at a national level. Early (within two years of index procedure) implant revision is a reasonable time window to examine the relationship between patient residence/travel and surgical outcomes, as a balance between the stability of the all-cause revision outcome and the time-varying nature of patient residence is required, with up to 15% of the Australian population changing address per year, and over 10% of those that do change residence, moving interstate (Australian Bureau of Statistics, 8-Nov-2022). While aseptic mechanisms may occur at any period, the first two years after surgery are considered the greatest risk period for periprosthetic infections, with up to 70% occurring within this timeframe (Tande & Patel, 2014).

To address this gap, the aim of this study was to assess whether the incidence of all-cause revision at up to 2 years following primary hip or knee total joint arthroplasty varies with the remoteness of a person’s place of residence at the time of the primary procedure. It was hypothesised that patients residing in areas considered rural or remote are at significantly greater risk of all-cause revision surgery in their first two years post-surgery compared to patients from metropolitan areas when controlling for potential confounding factors.

## Methods

### Registry Background

The Australian Orthopaedic Association National Joint Replacement Registry (AOANJRR) commenced data collection on 01 September 1999, achieving complete national implementation by mid-2002. Since then it has collected data on 99.2% of THAs and total knee arthroplasties performed in Australia (Australian Orthopaedic Association National Joint Replacement Registry (AOANJRR). Hip, Knee & Shoulder Arthroplasty: 2022 Annual Report, 2022). These data are externally validated against patient-level data provided by all Australian state and territory health departments. A sequential, multilevel matching process is used to identify any missing data which are subsequently obtained by follow-up with the relevant hospital. Each month, in addition to internal validation and data quality checks, all primary procedures are linked to any subsequent revision involving the same patient, joint and side. Data are also matched bi-annually to the Australian National Death Index data to identify patients who have died. Data was available for the present analysis up to Dec 31 2022.

### Data Model

A directed-acyclic-graph (DAG) method was used to map the relationships between the outcome (2 year cumulative revision rate) and the primary exposure (patient residence remoteness) to identify potential confounders for inclusion into a model to address the aim of the study. A DAG is a non-parametric diagrammatic presentation of the assumed data-generating process for a set of variables in a specified context. Variables and their measurements are depicted as nodes (or vertices) connected by unidirectional arcs (or arrows; hence ‘directed’) depicting the hypothesised relationships between them (Tennant et al., 2021). Confounders are identified by an open path (all arrows between nodes point in the same direction) passing through a node between the exposure and the estimand. To reduce *confounding bias*, the confounders identified in the DAG can be included in a multivariable model as covariates (Tennant et al., 2021). In this study, a DAG was constructed using the browser-based version of Daggity v3.0 (Textor et al., 2016) (Figure 1).

**Figure 1:**
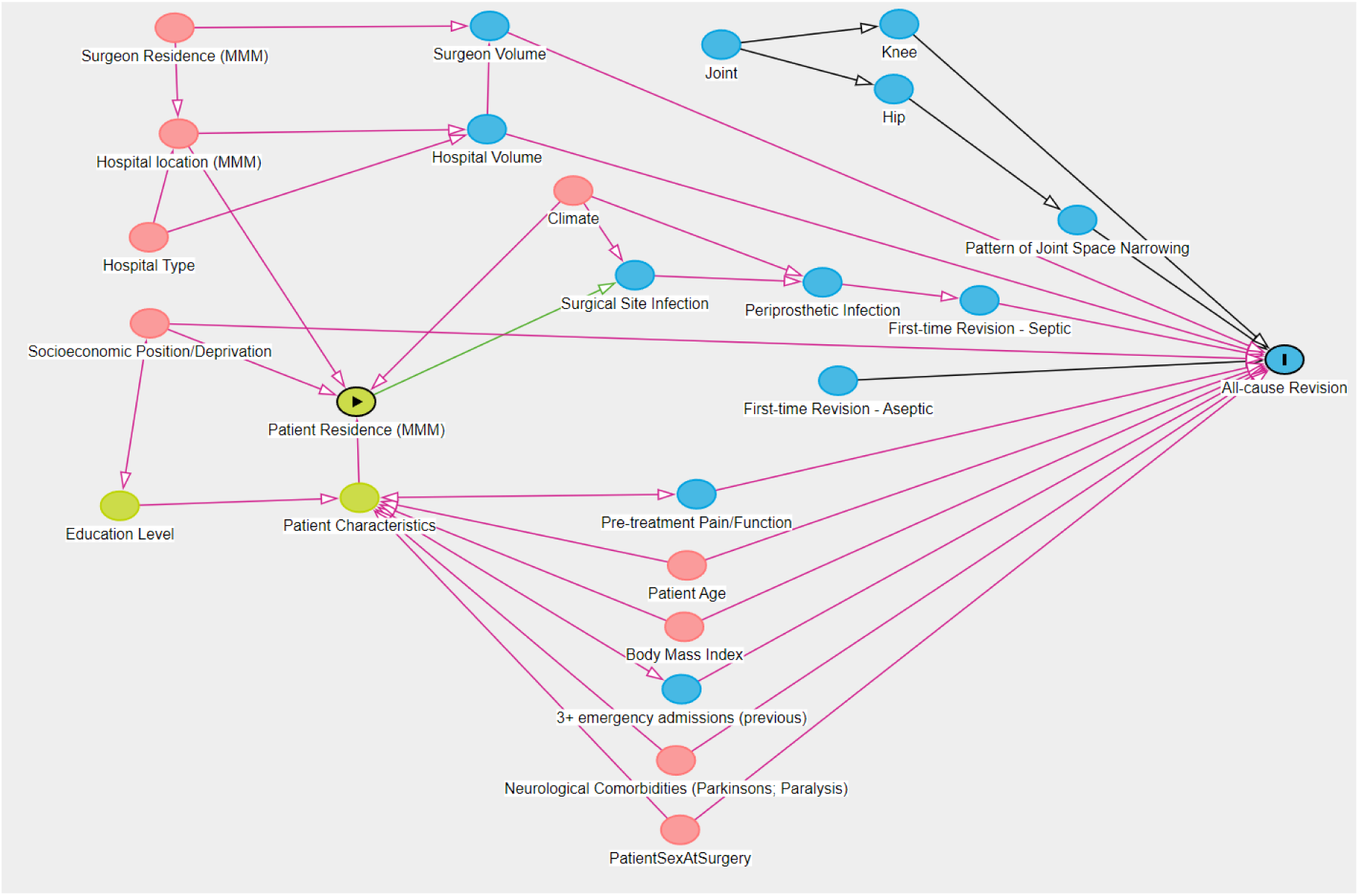
Directed acyclic graph of the relationship between patient residence, hospital location and all-cause revision in lower limb arthroplasty performed in Australia

### Primary Outcome

All cause revision within two years of the primary procedure.

### Primary Exposure (Patient Residence)

The patient residence was classified as part of the 2015 Modified Monash Model (Department of Health, 2019), which is structured in line with the Australian Standard Geographical Classification - Remoteness Area (ASGS-RA)(Australian Bureau of Statistics, 2021), which in turn relies on definitions provided by the Accessibility/ Remoteness Index of Australia (ARIA) developed by the Commonwealth Department of Health and Aged Care (DHAC) and the National Key Centre For Social Applications of GIS (GISCA)(*Accessibility/Remoteness Index of Australia (ARIA)*, n.d.). The MMM comprises 7 classifications, from 1 (metropolitan) through to 7 (Very Remote) as per Table 1.

**Table 1:** Modified Monash Model of remote area classification in Australia and its Territories.

| MMM | Definition |
| --- | --- |
| 1 | All areas categorised ASGC-RA1 (Major cities and inner-regional areas with an ARIA+ value $0 \leq 2.4$) |
| 2 | Areas categorised ASGS-RA 2 (ARIA+ value 2.4) and ASGS-RA 3 (ARIA+ value $2.4 \leq 5.92$) that are in, or within 20km road distance, of a town with a population greater than 50,000 |
| 3 | Areas categorised ASGS-RA 2 and ASGS-RA 3 that are not in MM 2 and are in, or within 15km road distance, of a town with a population between 15,000 and 50,000 |
| 4 | Areas categorised ASGS-RA 2 and ASGS-RA 3 that are not in MM 2 or MM 3 and are in, or within 10km road distance, of a town with a population between 5,000 and 15,000 |
| 5 | All other areas in ASGS-RA 2 and 3 |
| 6 | All areas categorised ASGS-RA 4 (ARIA+ value $5.92 \leq 10.53$) that are not on a populated island that is separated from the mainland in the ABS geography and is more than 5km offshore. Islands that have an MM 5 classification with a population of less than 1,000 |
| 7 | All other areas; that being ASGS-RA 5 (ARIA+ value $>10.53$) and areas on a populated island separated from the mainland in the ABS geography and is more than 5km offshore |

ABS - Australian Bureau of Statistics

### Confounder Variables

Adjustments for covariates were prioritised based on a narrative literature review to develop the DAG described in Appendix A.

### Patient selection

Included cases were restricted to primary diagnosis of osteoarthritis for both total knee and total hip arthroplasties. Bilateral status was not considered.

### Statistical Analysis

The Australian Orthopaedic Association National Joint Replacement Registry (AOANJRR) (Australian Orthopaedic Association National Joint Replacement Registry (AOANJRR). Hip, Knee & Shoulder Arthroplasty: 2022 Annual Report, 2022) was queried to assess the influence of patient residence remoteness on the cumulative percent revision (CPR) rate up to 2 years after the primary procedure. An analysis of all procedures from the Registry inception to 31 December 2022 was performed by the AOANJRR. Cox proportional hazards models were applied to the dataset to compare CPRs between *different combinations of patient MMM and hospital MMM*. Hazard ratios were reported (with 95% CI and p-values) comparing between the different combinations. Patient residence MMM and hospital MMM were dichotomized into two classifications MMM 1-2 (metro-regional) and MMM 3-7 (rural-remote) to mitigate low samples in certain sub-groups. Cox proportional hazards models were adjusted for the following confounder patient characteristics:

● Age at time of surgery: Calculated as the difference between the date of birth and date of surgery. Included as a continuous covariate in the survival model.
● Sex: As listed on the data collection form at the time of surgery, with binary responses (male; female)
● IRSAD: The 2016 Index of Relative Socio-economic Advantage and Disadvantage summarises variables that indicate either relative advantage or disadvantage. This index ranks areas on a continuum from most disadvantaged to most advantaged (AUSTRALIAN BUREAU OF STATISTICS, 2016; Parkinson et al., 2018)
● Tropics: Each hospital included in the dataset was categorised into one of two geographic regions (tropic or temperate) as described by (Parkinson et al., 2018)

The assumption of proportional hazards was checked analytically for each model. If the interaction between the predictor and the log of time was statistically significant in the standard Cox model, then a time varying model was estimated. Time points were selected based on the greatest change in hazard, weighted by a function of events. Time points were iteratively chosen until the assumption of proportionality was met and HRs were calculated for each selected time-period. For the current study, if no time-period was specified, the HR was calculated over the entire follow-up period. All tests were two-tailed at 5% levels of significance. Statistical analysis was performed using SAS software version 9.4 (SAS Institute Inc., Cary, North Carolina).

## Results

### Sample characteristics

There were 842,479 primary TKA procedures with a diagnosis of osteoarthritis included in the analysis with 55.7% females, average age of 68.5±9.1 years, 10.6% with normal or underweight BMI classification and 93.3% presenting with ASA 2 or 3. The remoteness categories (MM) were broken down into 6 of the 7 available sub-categories with no patients residing in MM7 undergoing TKA. A considerable proportion of all cases were performed on patients residing in an MM1 locality (Table 2).

**Table 2A:** Location breakdown for patient residence and hospital for total knee arthroplasty.

| Variable | Patient Locality | Hospital Locality |
| --- | --- | --- |
| **Follow-Up Years** |  |  |
| Mean ± SD | 7.3 ± 5.1 | 7.3 ± 5.1 |
| Median (IQR) | 6.5 (3.1, 10.8) | 6.5 (3.1, 10.8) |
| Minimum | 0 | 0 |
| Maximum | 23.3 | 23.3 |
| **Age** |  |  |
| Mean ± SD | 68.5 ± 9.1 | 68.5 ± 9.1 |
| Median (IQR) | 69 (62, 75) | 69 (62, 75) |
| **Age Group** |  |  |
| <55 | 54,902 (6.5%) | 54,902 (6.5%) |
| 55-64 | 222,341 (26.4%) | 222,341 (26.4%) |
| 65-74 | 336,543 (39.9%) | 336,543 (39.9%) |
| ≥75 | 228,693 (27.1%) | 228,693 (27.1%) |
| **Gender** |  |  |
| Male | 373,189 (44.3%) | 373,189 (44.3%) |
| Female | 469,290 (55.7%) | 469,290 (55.7%) |
| **Patient MMM** |  |  |
| 1 | 519,714 (61.7%) | 629,426 (74.7%) |
| 2 | 84,706 (10.1%) | 98,232 (11.7%) |
| 3 | 76,032 (9.0%) | 101,636 (12.1%) |
| 4 | 49,376 (5.9%) | 8,543 (1.0%) |
| 5 | 98,893 (11.7%) | 4,354 (0.5%) |
| 6 | 9,665 (1.1%) | 288 (0.0%) |
| 7 | 4,093 (0.5%) | 0 |
| **Femoral Cement** |  |  |
| Cementless | 336,476 (39.9%) | 336,476 (39.9%) |
| Cemented | 506,003 (60.1%) | 506,003 (60.1%) |
| **ASA Score<sup>a</sup>** |  |  |
| 1 | 28,042 (5.6%) | 28,042 (5.6%) |
| 2 | 271,110 (54.3%) | 271,110 (54.3%) |
| 3 | 194,549 (39%) | 194,549 (39%) |
| 4 | 5,193 (1%) | 5,193 (1%) |
| 5 | 14 (0%) | 14 (0%) |
| **BMI Category<sup>b</sup>** |  |  |
| Underweight (<18.50) | 732 (0.2%) | 732 (0.2%) |
| Normal (18.50-24.99) | 42,501 (10.4%) | 42,501 (10.4%) |
| Pre Obese (25.00-29.99) | 127,451 (31.3%) | 127,451 (31.3%) |
| Obese Class 1 (30.00-34.99) | 126,079 (30.9%) | 126,079 (30.9%) |
| Obese Class 2 (35.00-39.99) | 69,027 (16.9%) | 69,027 (16.9%) |
| Obese Class 3 ($\geq 40.00$) | 41,979 (10.3%) | 41,979 (10.3%) |
| **TOTAL** | **842,479** | **842,479** |

Abbreviations: SD - standard deviation, IQR - interquartile range, ASA - American Society of Anesthesiologists, BMI - Body Mass Index (kg/m<sup>2</sup>)
<sup>a</sup>Excludes 343,571 procedures with unknown ASA score
<sup>b</sup>Excludes 434,710 procedures with unknown BMI category

**Table 2B:** Location breakdown for patient residence and hospital for total hip arthroplasty.

| Variable | Patient Locality | Hospital Locality |
| --- | --- | --- |
| **Follow-Up Years** |  |  |
| Mean ± SD | 7.2 ± 5.2 | 7.2 ± 5.2 |
| Median (IQR) | 6.3 (3, 10.6) | 6.3 (3, 10.6) |
| Minimum | 0 | 0 |
| Maximum | 23.3 | 23.3 |
| **Age** |  |  |
| Mean ± SD | 68.2 ± 10.7 | 68.2 ± 10.7 |
| Median (IQR) | 69 (61, 76) | 69 (61, 76) |
| **Age Group** |  |  |
| <55 | 55,383 (10.7%) | 55,383 (10.7%) |
| 55-64 | 123,492 (23.8%) | 123,492 (23.8%) |
| 65-74 | 186,097 (35.9%) | 186,097 (35.9%) |
| $\geq 75$ | 153,499 (29.6%) | 153,499 (29.6%) |
| **Gender** |  |  |
| Male | 238,642 (46%) | 238,642 (46%) |
| Female | 279,829 (54%) | 279,829 (54%) |
| **Patient MMM** |  |  |
| 1 | 323,036 (62.3%) | 392,849 (75.8%) |
| 2 | 50,772 (9.8%) | 56,060 (10.8%) |
| 3 | 46,126 (8.9%) | 60,519 (11.7%) |
| 4 | 28,670 (5.5%) | 6,134 (1.2%) |
| 5 | 62,405 (12.0%) | 2,750 (0.5%) |
| 6 | 5,542 (1.1%) | 159 (0.0%) |
| 7 | 1,920 (0.4%) | 0 (0%) |
| **Femoral Cement** |  |  |
| Cementless | 323,921 (62.5%) | 323,921 (62.5%) |
| Cemented | 194,550 (37.5%) | 194,550 (37.5%) |
| **ASA Score<sup>a</sup>** |  |  |
| 1 | 27,352 (8.8%) | 27,352 (8.8%) |
| 2 | 167,419 (54.1%) | 167,419 (54.1%) |
| 3 | 110,395 (35.7%) | 110,395 (35.7%) |
| 4 | 4,336 (1.4%) | 4,336 (1.4%) |
| 5 | 18 (0%) | 18 (0%) |
| **BMI Category<sup>b</sup>** |  |  |
| Underweight (<18.50) | 1,835 (0.7%) | 1,835 (0.7%) |
| Normal (18.50-24.99) | 52,397 (20.6%) | 52,397 (20.6%) |
| Pre Obese (25.00-29.99) | 93,380 (36.7%) | 93,380 (36.7%) |
| Obese Class 1 (30.00-34.99) | 65,292 (25.7%) | 65,292 (25.7%) |
| Obese Class 2 (35.00-39.99) | 27,745 (10.9%) | 27,745 (10.9%) |
| Obese Class 3 (≥40.00) | 13,500 (5.3%) | 13,500 (5.3%) |
| **TOTAL** | **518,471** | **518,471** |

Abbreviations: SD - standard deviation, IQR - interquartile range, ASA - American Society of Anesthesiologists, BMI - Body Mass Index (kg/m<sup>2</sup>)
<sup>a</sup>Excludes 208,951 procedures with unknown ASA score
<sup>b</sup>Excludes 264,322 procedures with unknown BMI category

There were 518,471 primary THA procedures undertaken for osteoarthritis included in the analysis with 54.0% females, average age of 68.2±10.7 years, 21.3% with normal or underweight BMI classification and 89.8% presenting with ASA 2 or 3. Numbers revised can be found in Tables 3A and 3B.

### Cumulative Percent Revision

For TKR procedures, the overall 2 year CPR ranged from 1.8% (95% CI 1.7 - 1.9) for MM3-7 patients receiving an arthroplasty in a MM3-7 hospital, to 2.2% (95% CI 1.8 - 2.7) for MM1-2 patients travelling to a MM3-7 hospital (Table 3A). Patients residing in MM3-7 areas that travelled to a hospital within a metropolitan area or regional centre (MM1-2) had a significantly higher rate of revision following TKA compared to patients that were treated at a hospital MM 3-7 (HR: 1.11, 95% CI 1.05 - 1.18, P = 0.001) within 2 year follow-up of the primary procedure (Figure 2). Patients residing in MM3-7 areas that stayed in these areas for their operation displayed a significantly reduced revision rate compared to metropolitan patients (MM1-2) that stayed in-area (hospital MM1-2) for their joint replacement (HR: 0.90, 95% CI 0.85 - 0.95, P <0.001). Infection was the dominant reason for TKA revision for patients in the follow-up period, particularly for those residing in MM1-2 and receiving an operation from a hospital in an area MM3-7 (Figure 3).

**Figure 2:**
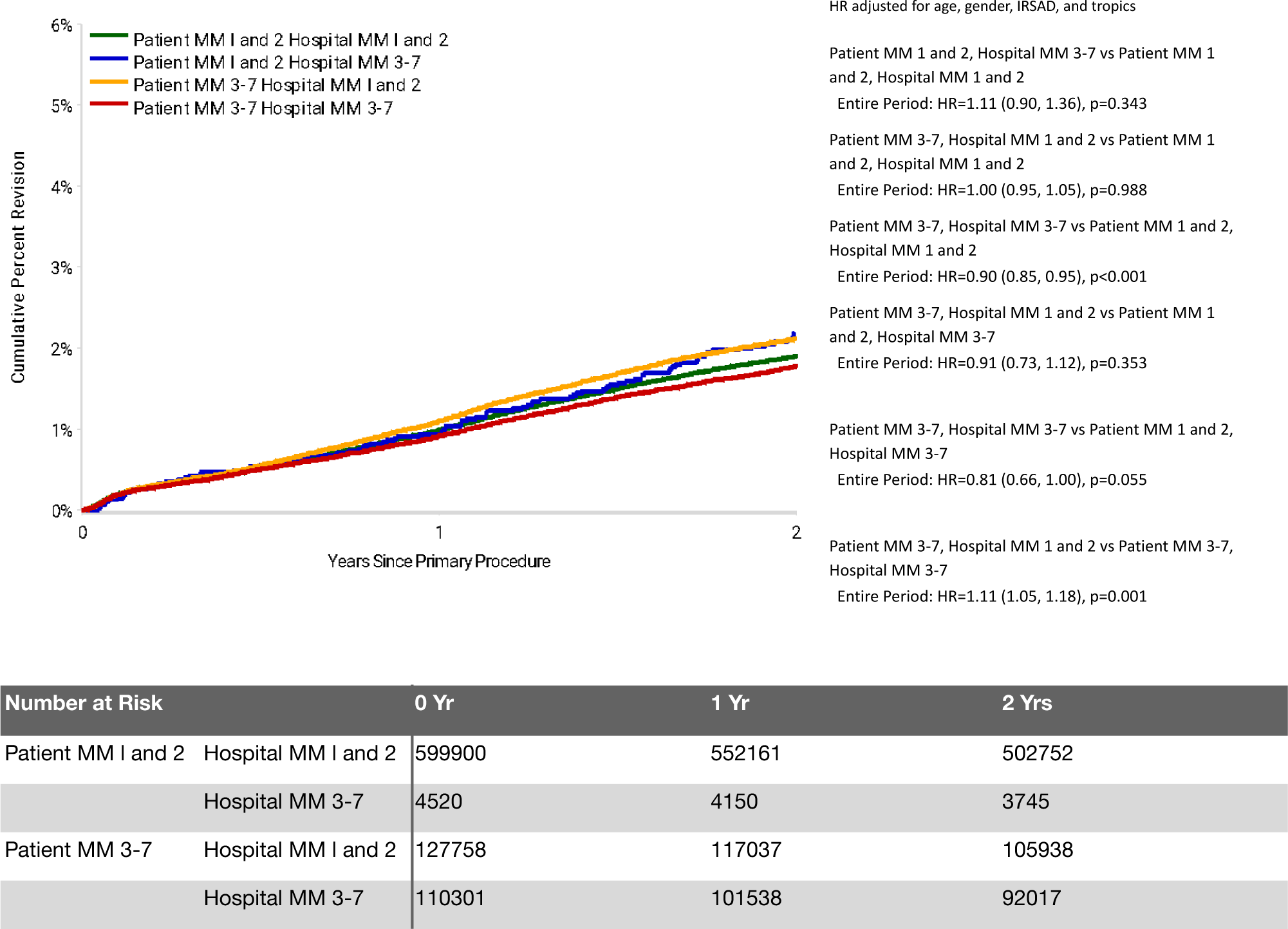
Cumulative Percent Revision of Primary Total Knee Replacement by Patient Locality Classification (Primary Diagnosis OA, Revision within 2 Years)

**Figure 3:**
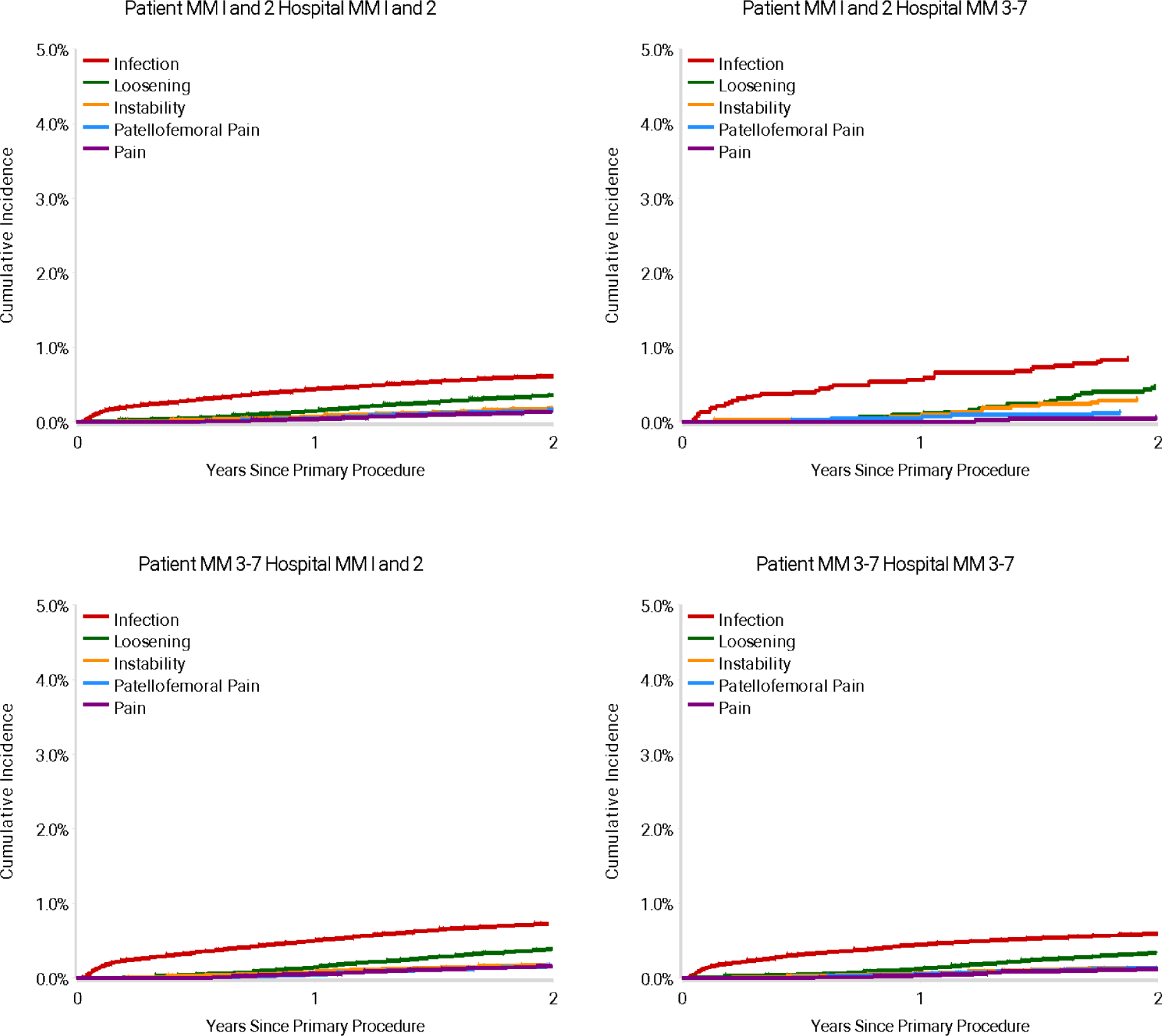
Cumulative Incidence Revision Diagnosis of Primary Total Knee Replacement by Patient and Hospital Locality (Primary Diagnosis OA, Revision within 2 Years)

For THR cases with diagnosis of osteoarthritis, overall 2 year cumulative revision rates ranged from 2.1% (95% CI 1.6 - 2.7) for MM1-2 patients receiving an arthroplasty in a MM3-7 hospital, to 2.1% (95% CI 2.0 - 2.2) for all other patient-hospital subgroups. There was no significant statistical difference in the rate of revision between any of the patient and hospital combinations for total hip replacement (Table 3B, Figure 4). While infection was a dominant reason for THA revision for patients in the follow-up period, this pattern was not as distinct as for TKA, with prosthesis dislocation or instability a prominent reason for revision in patients travelling “out of area” (top right and bottom left panels of Figure 5).

**Figure 4:**
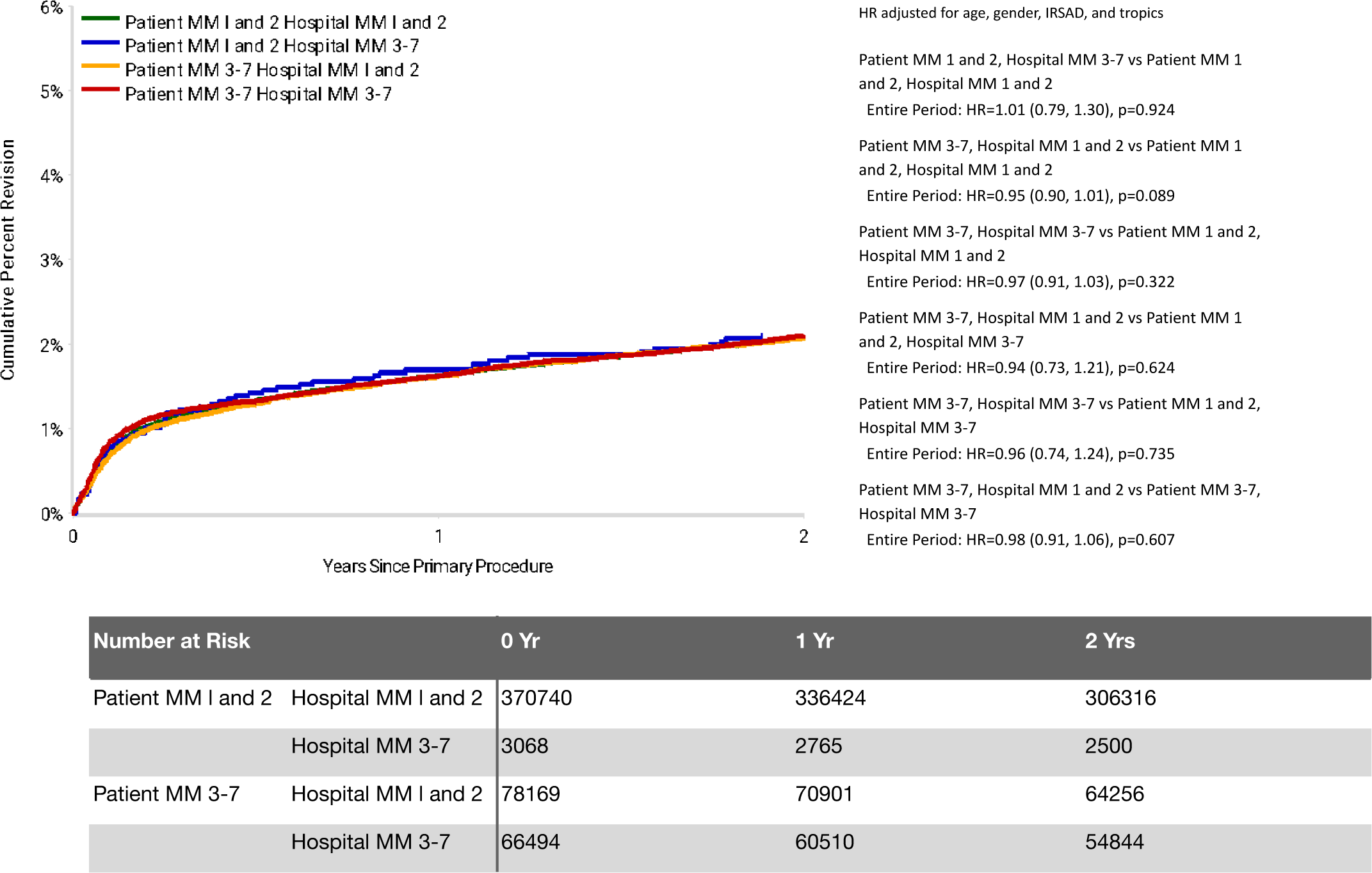
Cumulative Percent Revision of Primary Total Conventional Hip Replacement by Patient Locality Classification (Primary Diagnosis OA, Revision within 2 Years)

**Figure 5:**
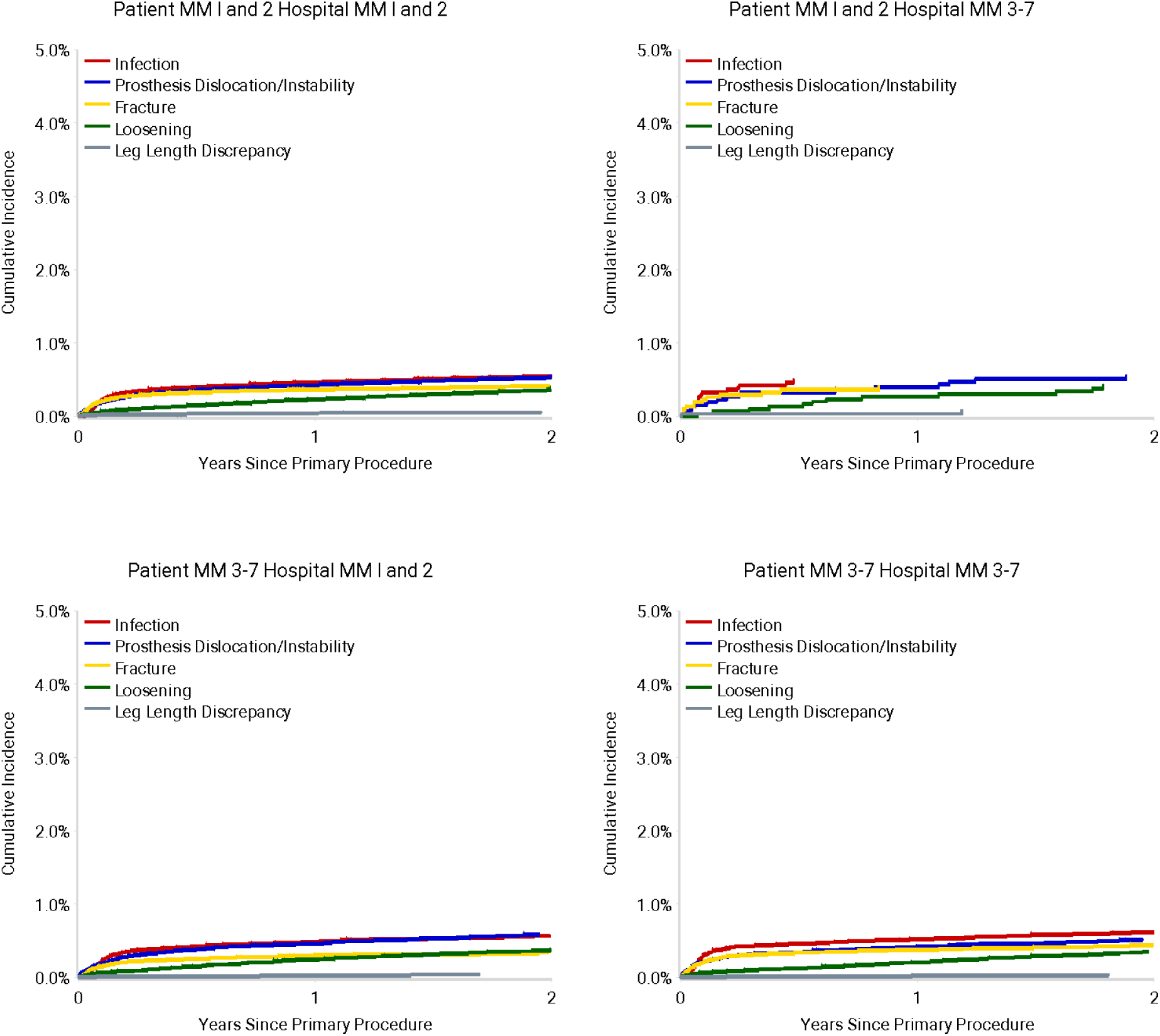
Cumulative Incidence Revision Diagnosis of Primary Total Hip Replacement by Patient and Hospital Locality (Primary Diagnosis OA, Revision within 2 Years)

**Table 3A:** Yearly Cumulative Percent Revision of Primary Total Knee Replacement by Patient Remoteness Classification (Primary Diagnosis OA, Revision within 2 Years)

| Patient MMM | Hospital MMM | N Revised | N Total | 1 Yr | 2 Yrs |
| --- | --- | --- | --- | --- | --- |
| Patient MM 1 and 2 | Hospital MM 1 and 2 | 10637 | 599900 | 1.0 (1.0, 1.0) | 1.9 (1.9, 1.9) |
|  | Hospital MM 3-7 | 91 | 4520 | 0.9 (0.7, 1.3) | 2.2 (1.8, 2.7) |
| Patient MM 3-7 | Hospital MM 1 and 2 | 2514 | 127758 | 1.1 (1.0, 1.2) | 2.1 (2.0, 2.2) |
|  | Hospital MM 3-7 | 1824 | 110301 | 0.9 (0.9, 1.0) | 1.8 (1.7, 1.9) |
| **TOTAL** |  | **15066** | **842479** |  |  |

**Table 3B:** Yearly Cumulative Percent Revision of Primary Total Hip Replacement by Patient Remoteness Classification (Primary Diagnosis OA, Revision within 2 Years)

| Patient MMM | Hospital MMM | N Revised | N Total | 1 Yr | 2 Yrs |
| --- | --- | --- | --- | --- | --- |
| Patient MM I and 2 | Hospital MM I and 2 | 7357 | 370740 | 1.6 (1.6, 1.7) | 2.1 (2.0, 2.1) |
|  | Hospital MM 3-7 | 62 | 3068 | 1.7 (1.3, 2.2) | 2.1 (1.6, 2.7) |
| Patient MM 3-7 | Hospital MM I and 2 | 1550 | 78169 | 1.6 (1.5, 1.7) | 2.1 (2.0, 2.2) |
|  | Hospital MM 3-7 | 1336 | 66494 | 1.6 (1.5, 1.7) | 2.1 (2.0, 2.2) |
| **TOTAL** |  | **10305** | **518471** |  |  |

## Discussion

The aim of this study was to assess whether the incidence of all-cause revision at up to 2 years following primary hip or knee total joint arthroplasty varies with the remoteness of a person’s place of residence at the time of the primary procedure. It was hypothesised that patients residing in areas considered rural or remote are at significantly greater risk of all-cause revision surgery compared to patients from metropolitan areas when controlling for potential confounding factors. A key finding from this analysis was that the hypothesis was not supported: patients that travelled out of area (rural/remote residence to metropolitan/regional hospital and vice versa) demonstrated significantly higher CPR after TKA compared to patients that stayed within the same remoteness category. Furthermore, patients residing in rural-remote areas that stayed in these areas for their operation displayed a significantly reduced revision rate compared to metro-regional patients that stayed in-area for their joint replacement. However, these findings were not replicated for THA and no significant differences in CPR were observed between remoteness combinations of patient residence and site of procedure.

The finding that non-metropolitan patients are at reduced risk of revision within two years of the primary procedure is counter-intuitive given the known geographic variation in health services. There are numerous elements of the outcome of joint arthroplasty (procedure revision) that depend on and are influenced by the reported deficiencies in health care provision in non-metropolitan areas. In short, for revision incidence to truly differ between areas, it must be assumed that there is equality in the patient sample with respect to: access to services and specialist care; diagnostic accuracy; access to timely revision surgery and its associated components (e.g. theatre and instrumentation availability, surgeon experience); and the availability of sufficient perioperative and post-operative services. The proportion of patients presenting with a symptomatic primary replacement and deciding that the risk-benefit favours surgery also needs to be equal between regions for the findings to hold true.

Given the reported deficiencies in health care provision in regional, rural and remote areas in Australia, there is a risk that the findings are an artefact of one or more of the assumptions failing to hold. For example, it could be that in cases of periprosthetic infection, patients returning to rural-remote areas and seeking treatment for subsequent infection are more likely to be offered implant retention and suppression with antibiotics (non-revision) than washout with implant exchange (revision) as a first-line treatment. This may be mitigated by travel distance, with patients travelling long distances to return after a primary procedure offered more aggressive reintervention options to reduce travel burden over time.

A key finding of this study is that TKA patients that travel out of their residential area regardless of remoteness category for their primary procedure have significantly higher CPR within the first two years after surgery, than those that stay. The reasons for this finding are not immediately apparent, however the issue of travel distance between residence and place of care is not new. A patient that has travelled out of area for the primary procedure with an issue during the postoperative period will i) present with an issue to the primary surgeon; ii) present to primary care/emergency with subsequent specialist referral; iii) receive local management, through peer-to-peer referral from i) or ii). Travel distance has been inversely associated with higher representation rates to the treating hospital for other types of surgery (Simpson et al., 2019),, while others have failed to detect an effect of travel distance on complication rates in reverse shoulder arthroplasty (Dubiel et al., 2022), although the analysis was hampered by referral bias at a single tertiary centre. Nevertheless, patients that travel large distances to access services for their primary procedure may be at greater risk of *care fragmentation* when re-presenting with a symptomatic arthroplasty, where care is delivered by different services to the same patient over time and with limited integration of clinical information. Care fragmentation is a common occurrence (∼25% of all readmissions), particularly in patients that travel long distances to access surgery (Tsai et al., 2015), although fragmented care may be less prevalent in orthopaedics in general (Kailasam et al., 2019). The findings of the present study may reflect that, in the sub-population of patients travelling out of residential areas for surgery, they are more likely to represent to a different health service, where a higher propensity to offer a revision procedure for a primary performed elsewhere may exist due to gaps created by poor integration of clinical information. Nevertheless, the aetiology of patients travelling out of area undergoing higher rates of revision for infection remains unexplored more broadly and further investigation is required to better understand the pathways for infection treatment across geographical areas.

While a relationship between patient remoteness and 2-year CPR was observed for TKA, this finding was not replicated for THA. The lack of replication in THA cases indicates that there may be a mediating effect of *joint* on the influence that patient remoteness has on early revision risk after lower limb arthroplasty. There are three possible mechanisms for this difference between hip and knee arthroplasty. The first is that the main indications for revision in THA were prosthesis dislocation/instability and infection compared to TKA, which were infection and component loosening. The influence of travel distance on management of prosthesis dislocation may be less pervasive than for infection, where the assumptions to observe differences in revision between remoteness regions may hold despite variations in the clinical management offered between clinics. The second mechanism that may explain the differences observed between joints is that the delay in diagnosis of infected arthroplasty is significantly shorter in hips compared to knees (25 vs 42 days) (Lewis et al., 2015). While the mechanisms of differences remain unresolved, the authors hypothesised that ambiguity of symptoms in infected knees compared to hips may contribute to delayed diagnosis. The third mechanism is that the distribution of primary cases across remoteness classifications differed between knees and hips. While the analysis technique used to assess CPR is relatively robust to variations in subgroup sample sizes; the differences in distribution may also reflect variations in service access. In particular, regional access to high-volume specialist care centres for THA, while for TKA this appears to be concentrated in MM1-2 localities based on the distribution of primary cases. Further work is required to elucidate the patterns of service access for symptomatic arthroplasties in relation to remoteness classification.

The present study provides new national-level data on the relationship between patient geographic residence and lower limb joint arthroplasty outcomes. However, the scope of interpretation should be shaped considering the limitations of the analysis. Firstly, the present analysis was constrained by the reduced sample sizes available in the non-metropolitan patients undergoing subsequent revision. For this reason, the MM classification, which is an ordinal variable, was collapsed into binary categories leading to reductions in the resolution and sensitivity of the exposure. Similarly, although local climate described by humidity and apparent temperature (continuous variables) has been associated with periprosthetic joint infection risk (Armit et al., 2018; Parkinson et al., 2018), in the present analysis this was included as a covariate by dichotomising MM categories into a binary category based on their relationship to the Tropic of Capricorn. This approach introduces a risk of differential classification bias (Lambert, 2011), incomplete confounder correction, as well as loss of analysis power (Naggara et al., 2011). Classification bias and spectrum bias (Lambert, 2011) may also be present in the revision indication, due to different levels of access to diagnostic services for patients in regional/rural communities, due to disparities in the health workforce (Legislative Council. Portfolio Committee No. 2 – Health, 2022; *Rural and Remote Health*, 2022). These biases have been addressed indirectly in earlier critiques of revision and survival as a registry endpoint (Goodfellow et al., 2010; Wylde & Blom, 2011). Future work should examine data linkage to health services utilisation (e.g. Medicare Benefits Schedule) pre-empting the shared decision to undergo a revision procedure. Secondly, while every attempt was made to generate unbiased estimates for patient remoteness using data linkage with other national datasets, the present findings remain at risk of unmeasured confounding, as any registry analysis (Stammers et al., 2017), but particularly a post-hoc analysis that is beyond the original concept of the dataset. The two most important confounders that should be considered in future research are the state/territory in which the primary procedure is performed and the presence of neurological comorbidities. The federated system of healthcare in Australia means that considerable variation in the availability of revision services may be attributed to the state/territory in which the primary procedure was performed, as well as being associated with the propensity for a patient to live in a rural/remote region. Further, it is plausible, but not well established, that the presence of a neurological comorbidity may factor into a person’s decision regarding their permanent place of residence. Services provided for those with neurological comorbidities (e.g. Parkinson’s disease) vary significantly between urban and regional areas within Australia (Lubomski et al., 2013). Other studies have demonstrated differences in prevalence in Parkinson’s disease across different geographical areas in both Australia (Queensland) (Peters et al., 2006), and the United Kingdom and is a risk factor for early revision in total hip and knee arthroplasty in the United Kingdom (Bottle et al., 2019).

## Conclusion

Travel distance, but not remoteness of a patient’s place of residence may be associated with cumulative risk of early revision (within 2 years) in TKA, particularly in regional-remote patients travelling out of area, but not for patients undergoing THA. Further work linking service utilisation prior to a revision procedure is required to clarify whether differences in revision are due to variable service provision or superior outcomes of the primary procedure. The present findings provide important guidance for national attempts to address variation across Australian states and territories with respect to regional arthroplasty services, with particular attention to the concentration of primary TKA in metropolitan centres. Further, future registry analyses should consider patient residence remoteness or travel distance as potential confounders for revision risk in other exposures, such as implant or technique selection.

## Data Availability

All data produced in the present study are available upon reasonable request to the authors.

## Acknowledgements

The authors acknowledge the efforts of Sophie Corfield, Meredith Harrison-Brown and Manaal Fatima in preparing the manuscript for submission.

## Authors’ contributions

MB: conceived the idea, contributed to the analysis request; reviewed and approved the manuscript.

CS: developed the idea; prepared the analysis request; developed the methods; liaised with the NJRR; prepared and revised the manuscript

CH: prepared the dataset; developed and implemented the methods; prepared and produced the analysis report; revised the manuscript

CV: Represents the NJRR; revised the manuscript

## References

1. Accessibility/Remoteness Index of Australia (ARIA). (n.d.). Hugo Centre for Population and Migration Studies | University of Adelaide. Retrieved October 11, 2021, from https://able.adelaide.edu.au/hugo-centre/services/aria

2. Ackerman, I. N., Bohensky, M. A., Zomer, E., Tacey, M., Gorelik, A., Brand, C. A., & de Steiger, R. (2019). The projected burden of primary total knee and hip replacement for osteoarthritis in Australia to the year 2030. BMC Musculoskeletal Disorders, 20(1), 90.

3. Afshar, N., English, D. R., & Milne, R. L. (2019). Rural-urban residence and cancer survival in high-income countries: A systematic review. Cancer, 125(13), 2172–2184.

4. Alvarez, P. M., McKeon, J. F., Spitzer, A. I., Krueger, C. A., Pigott, M., Li, M., & Vajapey, S. P. (2022). Socioeconomic factors affecting outcomes in total knee and hip arthroplasty: a systematic review on healthcare disparities. Arthroplasty (London, England), 4(1), 36.

5. Armit, D., Vickers, M., Parr, A., Van Rosendal, S., Trott, N., Gunasena, R., & Parkinson, B. (2018). Humidity a potential risk factor for prosthetic joint infection in a tropical Australian hospital. ANZ Journal of Surgery, 88(12), 1298–1301.

6. Australian Bureau of Statistics. (8-Nov-2022). Population movement in Australia. Australian Bureau of Statistics. https://www.abs.gov.au/articles/population-movement-australia

7. AUSTRALIAN BUREAU OF STATISTICS. (2016). Technical Paper: Socio-Economic Indexes for Areas (SEIFA) (No. 2033.0.55.001). Australian Bureau of Statistics. https://www.ausstats.abs.gov.au/ausstats/subscriber.nsf/0/756EE3DBEFA869EFCA258259000BA746/$File/SEIFA%202016%20Technical%20Paper.pdf

8. Australian Bureau of Statistics. (2021). Australian Statistical Geography Standard (ASGS) Edition 3: Vol. 5 - Remoteness structure (cat no. 1270.0.55.005). ABS.

9. Australian Orthopaedic Association National Joint Replacement Registry (AOANJRR). Hip, Knee & Shoulder Arthroplasty: 2022 Annual Report. (2022). AOA. https://aoanjrr.sahmri.com/documents/10180/732916/AOA+2022+AR+Digital/f63ed890-36d0-c4b3-2e0b-7b63e2071b16

10. Bottle, A., Parikh, S., Aylin, P., & Loeffler, M. (2019). Risk factors for early revision after total hip and knee arthroplasty: National observational study from a surgeon and population perspective. PloS One, 14(4), e0214855.

11. Brennan-Olsen, S. L., Vogrin, S., Graves, S., Holloway-Kew, K. L., Page, R. S., Sajjad, M. A., Kotowicz, M. A., Livingston, P. M., Khasraw, M., Hakkennes, S., Dunning, T. L., Brumby, S., Sutherland, A. G., Talevski, J., Green, D., Kelly, T.-L., Williams, L. J., & Pasco, J. A. (2019). Revision joint replacement surgeries of the hip and knee across geographic region and socioeconomic status in the western region of Victoria: a cross-sectional multilevel analysis of registry data. BMC Musculoskeletal Disorders, 20(1), 300.

12. Brennan-Olsen, S., Vogrin, S., Holloway, K. L., Page, R. S., Sajjad, M. A., Kotowicz, M. A., Livingston, P. M., Khasraw, M., Hakkennes, S., Dunning, T. L., Brumby, S., Pedler, D., Sutherland, A., Venkatesh, S., Williams, L. J., Duque, G., Graves, S., Lorimer, M., & Pasco, J. A. (2017). Geographic region, socioeconomic position and the utilisation of primary total joint replacement for hip or knee osteoarthritis across western Victoria: a cross-sectional multilevel study of the Australian Orthopaedic Association National Joint Replacement Registry. Archives of Osteoporosis, 12(1), 97.

13. Cosby, A. G., McDoom-Echebiri, M. M., James, W., Khandekar, H., Brown, W., & Hanna, H. L. (2019). Growth and Persistence of Place-Based Mortality in the United States: The Rural Mortality Penalty. American Journal of Public Health, 109(1), 155–162.

14. Department of Health. (2019). Modified Monash Model (MMM) 2015 [Dataset]. data.gov.au. https://researchdata.edu.au/modified-monash-model-mmm-2015/1433648

15. Dubiel, M. J., Kolz, J. M., Tagliero, A. J., Larson, D. R., Maradit Kremers, H., Cofield, R. R., Sperling, J. W., & Sanchez-Sotelo, J. (2022). Analysis of patient characteristics and outcomes related to distance traveled to a tertiary center for primary reverse shoulder arthroplasty. Archives of Orthopaedic and Traumatic Surgery. Archiv Fur Orthopadische Und Unfall-Chirurgie, 142(7), 1421–1428.

16. Goodfellow, J. W., O’Connor, J. J., & Murray, D. W. (2010). A critique of revision rate as an outcome measure: re-interpretation of knee joint registry data. The Journal of Bone and Joint Surgery. British Volume, 92-B(12), 1628–1631.

17. Harrington, R. A., Califf, R. M., Balamurugan, A., Brown, N., Benjamin, R. M., Braund, W. E., Hipp, J., Konig, M., Sanchez, E., & Joynt Maddox, K. E. (2020). Call to Action: Rural Health: A Presidential Advisory From the American Heart Association and American Stroke Association. Circulation, 141(10), e615–e644.

18. Ireland, M. J., March, S., Crawford-Williams, F., Cassimatis, M., Aitken, J. F., Hyde, M. K., Chambers, S. K., Sun, J., & Dunn, J. (2017). A systematic review of geographical differences in management and outcomes for colorectal cancer in Australia. BMC Cancer, 17(1), 95.

19. Kailasam, M., Guo, W., Hsann, Y. M., & Yang, K. S. (2019). Prevalence of care fragmentation among outpatients attending specialist clinics in a regional hospital in Singapore: a cross-sectional study. BMJ Open, 9(3), e022965.

20. Lambert, J. (2011). Statistics in brief: how to assess bias in clinical studies? Clinical Orthopaedics and Related Research, 469(6), 1794–1796.

21. Legislative Council. Portfolio Committee No. 2 – Health. (2022). Health outcomes and access to health and hospital services in rural, regional and remote New South Wales (No. 57; 57). New South Wales Parliament. https://www.parliament.nsw.gov.au/lcdocs/inquiries/2615/Report%20no%2057%20-%20PC%202%20-%20Health%20outcomes%20and%20access%20to%20services.pdf

22. Lewis, S. S., Dicks, K. V., Chen, L. F., Bolognesi, M. P., Anderson, D. J., Sexton, D. J., & Moehring, R. W. (2015). Delay in diagnosis of invasive surgical site infections following knee arthroplasty versus hip arthroplasty. Clinical Infectious Diseases: An Official Publication of the Infectious Diseases Society of America, 60(7), 990–996.

23. Long, H., Liu, Q., Yin, H., Wang, K., Diao, N., Zhang, Y., Lin, J., & Guo, A. (2022). Prevalence Trends of Site-Specific Osteoarthritis From 1990 to 2019: Findings From the Global Burden of Disease Study 2019. *Arthritis & Rheumatology (Hoboken*, N.J.), 74(7), 1172–1183.

24. Lubomski, M., Rushworth, R. L., Lee, W., Bertram, K., & Williams, D. R. (2013). A cross-sectional study of clinical management, and provision of health services and their utilisation, by patients with Parkinson’s disease in urban and regional Victoria. Journal of Clinical Neuroscience: Official Journal of the Neurosurgical Society of Australasia, 20(1), 102–106.

25. Naggara, O., Raymond, J., Guilbert, F., Roy, D., Weill, A., & Altman, D. G. (2011). Analysis by categorizing or dichotomizing continuous variables is inadvisable: an example from the natural history of unruptured aneurysms. AJNR. American Journal of Neuroradiology, 32(3), 437–440.

26. Parkinson, B., Armit, D., McEwen, P., Lorimer, M., & Harris, I. A. (2018). Is Climate Associated With Revision for Prosthetic Joint Infection After Primary TKA? Clinical Orthopaedics and Related Research, 476(6), 1200–1204.

27. Peters, C. M., Gartner, C. E., Silburn, P. A., & Mellick, G. D. (2006). Prevalence of Parkinson’s disease in metropolitan and rural Queensland: a general practice survey. Journal of Clinical Neuroscience: Official Journal of the Neurosurgical Society of Australasia, 13(3), 343–348.

28. Rural and remote health. (2022, July 7). Australian Institute of Health and Welfare. https://www.aihw.gov.au/reports/rural-remote-australians/rural-and-remote-health

29. Simpson, R. E., Wang, C. Y., House, M. G., Zyromski, N. J., Schmidt, C. M., Nakeeb, A., & Ceppa, E. P. (2019). Travel distance affects rates and reasons for inpatient visits after pancreatectomy. HPB: The Official Journal of the International Hepato Pancreato Biliary Association, 21(7), 818–826.

30. Stammers, J. G., Kuo, A., Hart, A. J., Smeeth, L., & Skinner, J. A. (2017). Registry Data-Valuable Lessons But Beware the Confounders. The Journal of Arthroplasty, 32(9S), S63–S67.

31. Tande, A. J., & Patel, R. (2014). Prosthetic joint infection. Clinical Microbiology Reviews, 27(2), 302–345.

32. Tennant, P. W. G., Murray, E. J., Arnold, K. F., Berrie, L., Fox, M. P., Gadd, S. C., Harrison, W. J., Keeble, C., Ranker, L. R., Textor, J., Tomova, G. D., Gilthorpe, M. S., & Ellison, G. T. H. (2021). Use of directed acyclic graphs (DAGs) to identify confounders in applied health research: review and recommendations. International Journal of Epidemiology, 50(2), 620–632.

33. Textor, J., van der Zander, B., Gilthorpe, M. S., Liskiewicz, M., & Ellison, G. T. (2016). Robust causal inference using directed acyclic graphs: the R package “dagitty.” International Journal of Epidemiology, 45(6), 1887–1894.

34. Theile, D. E., Philpot, S., Blake, M., Harrington, J., & Youl, P. H. (2019). Outcomes following colorectal cancer surgery: Results from a population-based study in Queensland, Australia, using quality indicators. Journal of Evaluation in Clinical Practice, 25(5), 834–842.

35. Tsai, T. C., Orav, E. J., & Jha, A. K. (2015). Care fragmentation in the postdischarge period: surgical readmissions, distance of travel, and postoperative mortality. JAMA Surgery, 150(1), 59–64.

36. Wylde, V., & Blom, A. W. (2011). The failure of survivorship. The Journal of Bone and Joint Surgery. British Volume, 93(5), 569–570.

