## Appendix A for "Stay or go? Outcomes of lower limb arthroplasty in patients travelling away from home for surgery: A cross-sectional analysis of the AOANJRR comparing patient residence and hospital remoteness"

### Justification of Confounder Variables

Age and key life events are important considerations and drivers of internal migration within Australia (Australian Government-Centre for Population, 2020). The relationship between age and risk of early revision appears relatively well established, with observations in recent NJRR reports (Australian Orthopaedic Association National Joint Replacement Registry (AOANJRR), 2020), a scoping review (Jasper et al., 2016), as well as analyses of registry data from the United States (Dy et al., 2014) and the United Kingdom (Bottle et al., 2019). Patient sex at the time of the primary procedure follows a similar pattern to age. Disparities between the proportion of males and females are marked across different areas of Australia and between Metropolitan and regional centres (*Regional Population by Age and Sex, 2020*, 2021). Similarly, differences in revision risk have been identified between sex for both hip and knee arthroplasty procedures. Patel et al. (2020) observed gender differences in outcomes of hip and knee arthroplasty in a survey of US based procedures between 2011 and 2017. Singh et al also reported differences in revision rates at 5 years between males and females in a study of procedures performed in Pennsylvania (USA) in 2002. A systematic review (Towle & Monnot, 2016) reported higher rates of revision in males following total hip arthroplasty. In contrast, body mass index (BMI) is likely a mediator rather than a confounder. While the relationship between body mass index and revision risk has been somewhat established in the contemporary literature for total hip and knee arthroplasty (Onggo et al., 2020, 2021), it is not clear whether patients with higher or lower BMI are attracted to living in certain areas, more likely is that the remoteness and availability of different services and infrastructure potentially shapes the BMI of its residents. The latter has been demonstrated with respect to a relationship between building density and resident BMI in the United States (Troy et al., 2018).

Climate as a confounder between the pathway from patient residence to all-cause revision through the mediators of surgical site infection and periprosthetic infection. The relationship between patient residence and climate from a causal perspective is not unidirectional. They are driven by each other with the climate related to where the patient residence is located relative to the equator as well as other major drivers of climate such as major oceans, mountain ranges and other geographical features. In Australia there are different climate regions of which North Queensland being an obvious case in point compared to Tasmania. Nevertheless, the climate of a region where an individual may decide to permanently reside can change and those temporal shifts are probably important to the risk of revision in regional areas over time. A number of studies in Australia have shown relationships between climate as defined by surrogate measures and the risk of surgical site infection as well as periprosthetic infection. A systematic review (Vickers et al., 2019) reported increased odds of post-operative infection for orthopaedic patients who undergo procedures during periods of higher apparent temperature. A more recent meta analysis also reported a higher rate of surgical site infections in areas with warm weather (Sahtoe et al., 2021). Humidity in particular has been identified as a risk factor for periprosthetic joint infection in regional areas in Australia and this was shown to be seasonal (Armit et al., 2018; Parkinson et al., 2018).

The influence of socioeconomic status (SES) or disadvantage on arthroplasty outcomes remains controversial. Some have failed to observe a relationship between household income and arthroplasty revision risk (Peltola & Järvelin, 2014) while others have noted that cancer survival is related to distance from treatment/service and not individual or area-based SES (Murchie et al., 2021). In contrast a Danish study reported increased mortality and revision rates in lower SES up to 1 year after total hip replacement (Edwards et al., 2021). In this study, SES was quantified with the Index of Relative Socio-economic Disadvantage (IRSD), which is a general socio-economic index that summarises a range of information about the economic and social conditions of people and households within an area (Australian Bureau of Statistics, 2018).

In this particular model, hospital location and those factors that determine where a hospital is planned and built are identified as confounders in this context. The causal mechanisms for where a hospital is located relative to where a patient resides and where a surgeon lives is not well documented in the literature. A hospital location is likely determined by the business case for either a public or private operated facility with different decision-making and planning processes. The decision to place a public hospital with capacity to perform joint arthroplasty is different from one that is established under a private operator model. Evidence to support this theory remains lacking. Surgeon volume is a known risk factor for early revision as is hospital volume (Kugler et al., 2021), which are interrelated concepts.

**References**

Armit, D., Vickers, M., Parr, A., Van Rosendal, S., Trott, N., Gunasena, R., & Parkinson, B. (2018). Humidity a potential risk factor for prosthetic joint infection in a tropical Australian hospital. *ANZ Journal of Surgery*, *88*(12), 1298–1301.

Australian Bureau of Statistics. (2018). *Main Features - IRSD*. ABS. https://www.abs.gov.au/ausstats/abs@.nsf/Lookup/by%20Subject/2033.0.55.001~2016~Main%20Features~IRSD~19

Australian Government-Centre for Population. (2020). *Why do people move? Understanding internal migration in Australia*. https://population.gov.au/docs/why_do_people_move_understanding_internal_migration_in_australia.pdf

Australian Orthopaedic Association National Joint Replacement Registry (AOANJRR). (2020). *2020 Hip, Knee & Shoulder Arthroplasty Annual Report*. AOA. https://aoanjrr.sahmri.com/documents/10180/689619/Hip%2C+Knee+%26+Shoulder+Arthroplasty+New/6a07a3b8-8767-06cf-9069-d165dc9baca7

Bottle, A., Parikh, S., Aylin, P., & Loeffler, M. (2019). Risk factors for early revision after total hip and knee arthroplasty: National observational study from a surgeon and population perspective. *PloS One*, *14*(4), e0214855.

Dy, C. J., Bozic, K. J., Pan, T. J., Wright, T. M., Padgett, D. E., & Lyman, S. (2014). Risk factors for early revision after total hip arthroplasty. *Arthritis Care & Research*, *66*(6), 907–915.

Edwards, N. M., Varnum, C., Overgaard, S., & Pedersen, A. B. (2021). Impact of socioeconomic status on the 90- and 365-day rate of revision and mortality after primary total hip arthroplasty: a cohort study based on 103,901 patients with osteoarthritis from national databases in Denmark. *Acta Orthopaedica*, *92*(5), 581–588.

Jasper, L. L., Jones, C. A., Mollins, J., Pohar, S. L., & Beaupre, L. A. (2016). Risk factors for revision of total knee arthroplasty: a scoping review. *BMC Musculoskeletal Disorders*, *17*, 182.

Kugler, C. M., Goossen, K., Rombey, T., De Santis, K. K., Mathes, T., Breuing, J., Hess, S., Burchard, R., & Pieper, D. (2021). Hospital volume-outcome relationship in total knee arthroplasty: a systematic review and dose-response meta-analysis. *Knee Surgery, Sports Traumatology, Arthroscopy: Official Journal of the ESSKA*. https://doi.org/10.1007/s00167-021-06692-8

Murchie, P., Fielding, S., Turner, M., Iversen, L., & Dibben, C. (2021). Is place or person more important in determining higher rural cancer mortality? A data-linkage study to compare individual versus area-based measures of deprivation. *International Journal of Population Data Science*, *6*(1), 1403.

Onggo, J. R., Ang, J. J. M., Onggo, J. D., de Steiger, R., & Hau, R. (2021). Greater risk of all-cause revisions and complications for obese patients in 3 106 381 total knee arthroplasties: a meta-analysis and systematic review. *ANZ Journal of Surgery*, *91*(11), 2308–2321.

Onggo, J. R., Onggo, J. D., de Steiger, R., & Hau, R. (2020). Greater risks of complications, infections, and revisions in the obese versus non-obese total hip arthroplasty population of 2,190,824 patients: a meta-analysis and systematic review. *Osteoarthritis and Cartilage / OARS, Osteoarthritis Research Society*, *28*(1), 31–44.

Parkinson, B., Armit, D., McEwen, P., Lorimer, M., & Harris, I. A. (2018). Is Climate Associated With Revision for Prosthetic Joint Infection After Primary TKA? *Clinical Orthopaedics and Related Research*, *476*(6), 1200–1204.

Patel, A. P., Gronbeck, C., Chambers, M., Harrington, M. A., & Halawi, M. J. (2020). Gender and Total Joint Arthroplasty: Variable Outcomes by Procedure Type. *Arthroplasty Today*, *6*(3), 517–520.

Peltola, M., & Järvelin, J. (2014). Association between household income and the outcome of arthroplasty: a register-based study of total hip and knee replacements. *Archives of Orthopaedic and Traumatic Surgery. Archiv Fur Orthopadische Und Unfall-Chirurgie*, *134*(12), 1767–1774.

*Regional population by age and sex, 2020*. (2021, March 9). Australian Bureau of Statistics. https://www.abs.gov.au/statistics/people/population/regional-population-age-and-sex/latest-release

Sahtoe, A. P. H., Duraku, L. S., van der Oest, M. J. W., Hundepool, C. A., de Kraker, M., Bode, L. G. M., & Zuidam, J. M. (2021). Warm Weather and Surgical Site Infections: A Meta-analysis. *Plastic and Reconstructive Surgery. Global Open*, *9*(7), e3705.

Towle, K. M., & Monnot, A. D. (2016). An Assessment of Gender-Specific Risk of Implant Revision After Primary Total Hip Arthroplasty: A Systematic Review and Meta-analysis. *The Journal of Arthroplasty*, *31*(12), 2941–2948.

Troy, A. R., Bonnell, L. N., & Littenberg, B. (2018). Relationship Between the Built Environment and Body Mass Index in a Rural Context: A Cross-Sectional Study from Vermont. *Cureus*, *10*(7), e3040.

Vickers, M. L., Pelecanos, A., Tran, M., Eriksson, L., Assoum, M., Harris, P. N., Jaiprakash, A., Parkinson, B., Dulhunty, J., & Crawford, R. W. (2019). Association between higher ambient temperature and orthopaedic infection rates: a systematic review and meta-analysis. *ANZ Journal of Surgery*, *89*(9), 1028–1034.
